# Associations of pet ownership and pet attachment with a metabolite-based psychosocial distress score and circulating metabolomic profiles

**DOI:** 10.64898/2026.09.03.26361932

**Authors:** Oana A. Zeleznik, Susanne Strohmaier, Phuong Anh Le, A. Heather Eliassen, Curtis Huttenhower, Carri Westgarth, Magdalena Żebrowska, Tianyi Huang, Francine Laden, Jaime Hart, Bernard Rosner, Ichiro Kawachi, Jorge Chavarro, Olivia I. Okereke, Eva S. Schernhammer

**Author notes:** **Corresponding author:** Eva S. Schernhammer, MD, DrPH Channing Division of Network Medicine, Brigham and Women’s Hospital and Harvard Medical School 181 Longwood Avenue, Boston, MA 02115,; Oana A. Zeleznik, PhD, Department of Population Health Sciences Weill Cornell Medicine, 425 East 61^st^ Street, New York, NY 10065.

## Abstract

**Introduction:** Pet ownership is widespread, with many owners reporting emotional benefits, but evidence on its psychological and biological effects remains mixed. Prior research has linked strong pet attachment, particularly to dogs, to lower psychosocial distress, but underlying biological mechanisms are not well understood. We examined associations of pet ownership and attachment with a metabolite-based psychosocial distress score (MDS) and broader metabolic profiles, distinguishing between dogs and cats.

**Methods:** We analyzed data from 213 participants (131 pet owners, 82 non-owners), using the Lexington Attachment to Pets Scale and a previously developed MDS reflecting psychosocial distress. Linear regression assessed associations between pet variables and MDS or individual metabolites, adjusting for demographic, lifestyle, and health factors. Metabolite set enrichment analysis identified associated metabolite classes.

**Results:** Pet ownership and pet attachment were not associated with MDS overall. However, stronger pet attachment was associated with lower MDS (β(95%CI)=-0.58(−1.07,-0.08), p=0.02) among dog owners but not cat owners. Across all measured metabolites, N6,N6-dimethyllysine was significantly associated with pet attachment. Several metabolite classes were associated with pet ownership and attachment, with some in opposite directions. For example, triglycerides with than three double bonds were positively associated with ownership but inversely with attachment, particularly among cat owners.

**Conclusion:** Although pet ownership and attachment were not associated with MDS overall, stronger attachment was associated with lower MDS among dog owners but not cat owners. Pet ownership and attachment showed distinct metabolomic patterns, with differences between dog and cat owners. Further studies are needed to replicate and expand on these findings.

## Introduction

Pet ownership is widespread across the world, with more than half of the global population owning pets. The highest rates are found in the US, Europe, and parts of Asia, where dogs and cats dominate household companionship. While 82% of owners cite benefits like companionship and stress relief [1], research on human-animal interactions remains inconclusive, ranging from benefits to no effect or harm [2, 3]. Moreover, studies on biological mechanisms of pet ownership and attachment are limited, complicating our understanding of human-animal interactions.

The metabolome links genotype and phenotype [4] and includes small molecules like nucleotides, amino acids, carbohydrates, fatty acids and lipids. Metabolite levels, shaped by genes, lifestyle, and environment, reflect downstream processes [5]. Thus, the systematic analysis of metabolites can uncover valuable insights into the biological processes underlying human-animal interactions. Metabolomic profiles offer a comprehensive view of an individual’s metabolic state and have been used to elucidate some of the biological pathways underlying several psychosocial factors [6–9].

Previously, we reported a statistically significant association between pet attachment, particularly attachment to dogs, but not pet ownership, and multiple questionnaire-based psychosocial distress measures [10]. Metabolomic profiles have revealed biological pathways linked to psychosocial factors [6–9], and we previously derived a metabolite-based psychosocial distress score (MDS) reflecting depression and anxiety, which is associated with cardiovascular disease risk [6, 11, 12]. Building on these findings, we conducted the first study to: 1) Assess the association of pet attachment and ownership, by pet type, with the previously derived MDS, thereby providing an orthogonal validation of our prior results showing that pet attachment, especially to dogs, is linked to psychosocial distress measures, whereas pet ownership alone is not; and 2) Identify more broad metabolic profiles, beyond those related to psychosocial measures, associated with pet ownership and attachment, distinguishing between cats and dogs.

## Methods

### Study population

In 2013, 688 participants in the Nurses’ Health Study II (NHS2) [13] - a longitudinal cohort of registered US nurses - were invited to take part in the Mind Body Study (MBS) [14]. Those who agreed to participate obtained an initial kit to collect blood, urine, hair, nail and saliva samples, and were asked to complete (twice, about 12 months apart) a comprehensive online questionnaire assessing psychosocial factors. Women who reported on childhood abuse were oversampled in the MBS to increase the proportion of individuals with psychosocial suffering. The repeated questionnaire was aimed to test consistency and reproducibility of assessed measures, which was confirmed by high intraclass correlation (ICCs = 0.51-0.81) [14]. The study protocol was approved by the Institutional Review Board of Brigham and Women’s Hospital and the Committee on the Use of Human Subjects in Research of Harvard T.H. Chan School of Public Health (Boston, MA, USA). Voluntary return of questionnaires indicates informed consent. The study was conducted in accordance with all relevant ethical guidelines and regulations, including the Declaration of Helsinki.

### Assessment of pet ownership, pet attachment and other measures

In the two online surveys, participants were asked if they owned any pets and which type. They could choose only one from six categories (dog, cat, bird, fish, other pets or no pets in household). In this study we use the information from the first questionnaire which was administered about one month after the blood sample collection. Those who reported owning a dog were defined as dog owners while those who reported owning a cat were defined as cat owners. Pet owners included dog and cat owners as well as those who reported owning fish or other unspecified pets. As non-pet owners we defined those who reported not owning pets. Pet owners were also asked six questions from the Lexington Attachment to Pets Scale (LAPS) [15]. They assessed the participants’ feelings and behaviors towards their pets, such as considering them a friend, talking to them, finding happiness in owning a pet, discussing their pet with others, playing with and considering their pet as a part of the family. Participants could respond with options: strongly disagree, disagree, agree, strongly agree or don’t know, which were assigned numerical values. The total LAPS score, which can range from 0 to 18, was calculated by summing up the responses, with higher scores indicating higher attachment. We used the inverse normal transformation to ensure a normal distribution of LAPS scores.

Psychological distress was defined earlier by our group in this study as a summary measure including 10-item Centre for Epidemiologic Studies Depression Scale, Kessler Psychological Distress Scale, 7-item Generalized Anxiety Disorder scale, Crown Crisp Experiential Index phobic anxiety subscale [10].

We considered age, race, body mass index (BMI), alcohol consumption (in g/day), physical activity (in METs hours/week), diet quality measured by the Alternative Healthy Eating Index (AHEI), smoking status, social-economic status, sleep quality, and fasting status at the time of blood collection as potential confounders.

### Metabolite profiling

Plasma metabolites were profiled at the Broad Institute of MIT and Harvard (Cambridge, MA) using three complementary liquid chromatography tandem mass spectrometry (LC-MS/MS) methods designed to measure polar metabolites and lipids as well as free fatty acids as described previously [16–19]. For each method, metabolite identities were confirmed using authentic reference standards or reference samples. Additional details are presented in the supplementary materials.

In total, 614 known metabolites were measured. Metabolites with a coefficient of variation (CV) >25% or an intraclass correlation coefficient (ICC) <0.4 among blinded QC samples (N=106), metabolites measured by multiple metabolomics methods (N=7), and metabolites missing in >25% of study participants (N=4) were excluded. Furthermore, metabolites with poor stability due to delayed processing were excluded (N=66) [19]. Among the remaining 430 metabolites (including amino acids, amino acids derivatives, amines, lipids, fatty acids, bile acids, etc.), N=349 exhibited good reproducibility within person over one to two years [19]. Most metabolites show reasonable stability within person over 10 years [20]. None of the 430 metabolites included in this study had missing values among the study participants.

### Statistical methods

#### Metabolite-based distress score

We estimated metabolic dysregulation related to psychosocial distress with a metabolite based distress score (MDS) derived by Balasubramanian et al. [11] in a case-control dataset nested within the Nurses’ Health Study consisting of 279 women with prevalent chronic distress (characterized by recurring experiences of high levels of depression and anxiety) and 279 matched controls. A total of 20 metabolites were selected into the MDS [11], using coefficients from a multi-variable logistic regression including all metabolites (Supplementary Table 1). Due to the metabolomic assays used in this study, our dataset included 19 of the 20 MDS metabolites. In our study, the MDS is calculated as the linear combination of the 19 selected metabolites weighted by the coefficients estimated in the multi-variable linear regression [11].

MDS and metabolite levels were inverse-normal transformed for all analyses and modeled as continuous outcomes. Pet ownership (yes/no) was treated as categorical, and inverse-normal transformed pet attachment as continuous; both were considered exposures.

We used linear regression to estimate the mean difference in MDS levels as well as in the 19 MDS-metabolite levels and corresponding 95% confidence intervals (CI) comparing pet owners to those who do not own a pet. We used three nested statistical models: Model 1 adjusted for age. Model 2 additionally adjusted for date of, fasting status at, and menopausal status at the time of blood collection, and race. Model 3 additionally adjusted for body mass index, physical activity, Alternative Healthy Eating Index without alcohol, alcohol consumption, subjective sleep quality, and socio-economic status. In stratified analyses, we estimated mean difference in metabolite levels and corresponding 95% CI comparing cat owners or dog owners to those who do not own a pet.

We used linear regression to estimate the mean increase in MDS as well as the 19 MDS-metabolite levels and corresponding 95% CI for a one standard deviation (SD) increase in the transformed Lexington Attachment to Pets Scale. Transformed metabolite levels and the MDS were modeled continuously and represented the outcome while pet attachment was modeled continuously and represented the exposure. Similar to the pet ownership analyses, we used three nested statistical models. In stratified analyses, we estimated the mean increase in metabolite levels and corresponding 95% CI for a one standard deviation (SD) increase in the transformed Lexington Attachment to Pets Scale among cat owners and among dog owners.

Because the associations between MDS and the 19 MDS-metabolites with pet ownership and attachment represent *a priori* formulated hypotheses, we used a nominal p-value<0.05 to estimate statistical significance.

We used statistical mediation analysis to assess the proportion of the association of pet ownership or pet attachment with self-reported psychological distress explained by MDS [21, 22].

### Metabolic profiles associated with pet ownership and attachment

We used linear regression to estimate the mean difference in metabolite levels (N=430 metabolites) and corresponding 95% confidence intervals (CI) comparing pet owners to those who do not own a pet. Transformed metabolite levels were modeled continuously and represented the outcome while pet ownership was modeled as a categorical variable (yes/no) represented the exposure. Similar to the MDS-focused analyses, we used three nested statistical models. In stratified analyses, we estimated mean difference in metabolite levels and corresponding 95% CI comparing cat owners or dog owners to those who do not own a pet.

We used linear regression to estimate the mean increase in metabolite levels (N=430 metabolites) and corresponding 95% CI for a one standard deviation (SD) increase in the transformed Lexington Attachment to Pets Scale. Transformed metabolite levels were modeled continuously and represented the outcome while pet attachment was modeled continuously and represented the exposure. Similar to the pet ownership analyses, we used three nested statistical models. In stratified analyses, we estimated the mean increase in metabolite levels and corresponding 95% CI for a one standard deviation (SD) increase in the transformed Lexington Attachment to Pets Scale among cat owners and among dog owners.

We used the Metabolite Set Enrichment Analysis to estimate a normalized enrichment score (NES) and identify metabolite classes enriched among metabolites positively or inversely associated with pet ownership and pet attachment. We refer to metabolite classes enriched among positively associated metabolites as positively associated. We refer to metabolite classes enriched among inversely associated metabolites as inversely associated.

We used the number of effective tests (NEF) to account for testing multiple correlated hypotheses when investigating individual metabolites and the false discovery rate (FDR) when investigating metabolite classes. We used a NEF-corrected p value (NEF-p)<0.05 and FDR<0.05 to estimate statistical significance for individual metabolites and metabolite classes, respectively.

## Results

### 1. Participant characteristics

Our study included 213 participants with metabolomics data who provided information about their pets (Table 1). Among all participants, 82 reported not owning while 131 reported owning a pet. Pet owners included 76 dog, 52 cat, 1 fish, and 2 unspecified pet owners. Pet attachment (mean (standard deviation) = 15.36 (3.29)) was high among pet owners and similar for dog (15.54 (3.38)) and cat owners (15.31 (2.85)). Likely due to differences in socio-economic status, participant characteristics were somewhat different when comparing pet owners to those without pets. As expect, pet owners reported a slightly higher BMI and subjective sleep quality as well as lower physical activity, AHEI score excluding alcohol, alcohol consumption, compared to those without pets. Pet owners also included more current and past smokers and less never smokers compared to those not owning a pet.

**Table 1.** MBS study participant characteristics at the time of sample collection.

| Characteristics | Non pet owner | Pet owners* | Dog owners | Cat owners |
| --- | --- | --- | --- | --- |
| n | 82 | 131 | 76 | 52 |
| LAPS | NA | 15.36 (3.29) | 15.54 (3.38) | 15.31 (2.85) |
| Age, years | 61.02 (3.81) | 60.12 (4.07) | 59.85 (3.85) | 60.66 (4.21) |
| Not fasting, % | 12 (14.8) | 24 (18.6) | 16 ( 21.6) | 8 (15.4) |
| White Race, % | 77 (93.9) | 126 (96.2) | 74 ( 97.4) | 49 (94.2) |
| BMI, kg/m<sup>2</sup> | 25.47 (4.30) | 27.68 (6.80) | 27.83 (6.73) | 27.68 (7.07) |
| Physical activity, METs h/week | 34.19 (30.57) | 25.90 (25.24) | 27.28 (26.34) | 21.82 (21.84) |
| AHEI score excluding alcohol | 64.90 (12.88) | 62.95 (12.53) | 63.58 (12.05) | 61.95 (13.22) |
| Alcohol consumption, g/day | 8.76 (11.43) | 7.34 (11.23) | 8.81 (13.33) | 5.62 (7.43) |
| Smoking status (%) |  |  |  |  |
| never | 62 (75.6) | 84 (64.1) | 47 ( 61.8) | 35 (67.3) |
| past | 20 (24.4) | 43 (32.8) | 28 ( 36.8) | 15 (28.8) |
| current | 0 ( 0.0) | 4 ( 3.1) | 1 ( 1.3) | 2 ( 3.8) |
| Social-economic status | 0.69 (4.36) | 0.53 (4.22) | 0.79 (4.41) | 0.03 (3.93) |
| Subjective Sleep Quality | 0.74 (0.66) | 0.91 (0.60) | 0.88 (0.56) | 0.96 (0.66) |

Presented values represent means and standard deviations or percentages
\* Pets include 76 dogs, 52 cats, 1 fish and 2 unspecified pets.
LAPS, Lexington Attachment to Pets Scale
AHEI, Alternative Healthy Eating Index

### 2. Pet ownership, pet attachment, and the metabolite-based distress score (MDS)

Pet ownership (mean difference in MDS levels between pet owners and those not owning pets based on model 3 β (95% CI) = 0.11 (−0.24; 0.47), p = 0.53), dog ownership (model 3 β (95% CI) = 0.17 (−0.26; 0.60), p = 0.44) and cat ownership (model 3 β (95% CI) = 0.07 (−0.34; 0.48), p = 0.74) were not associated with MDS in either model (Figure 1, Supplementary Figure 1, and Supplementary Table 1).

**Figure 1:**
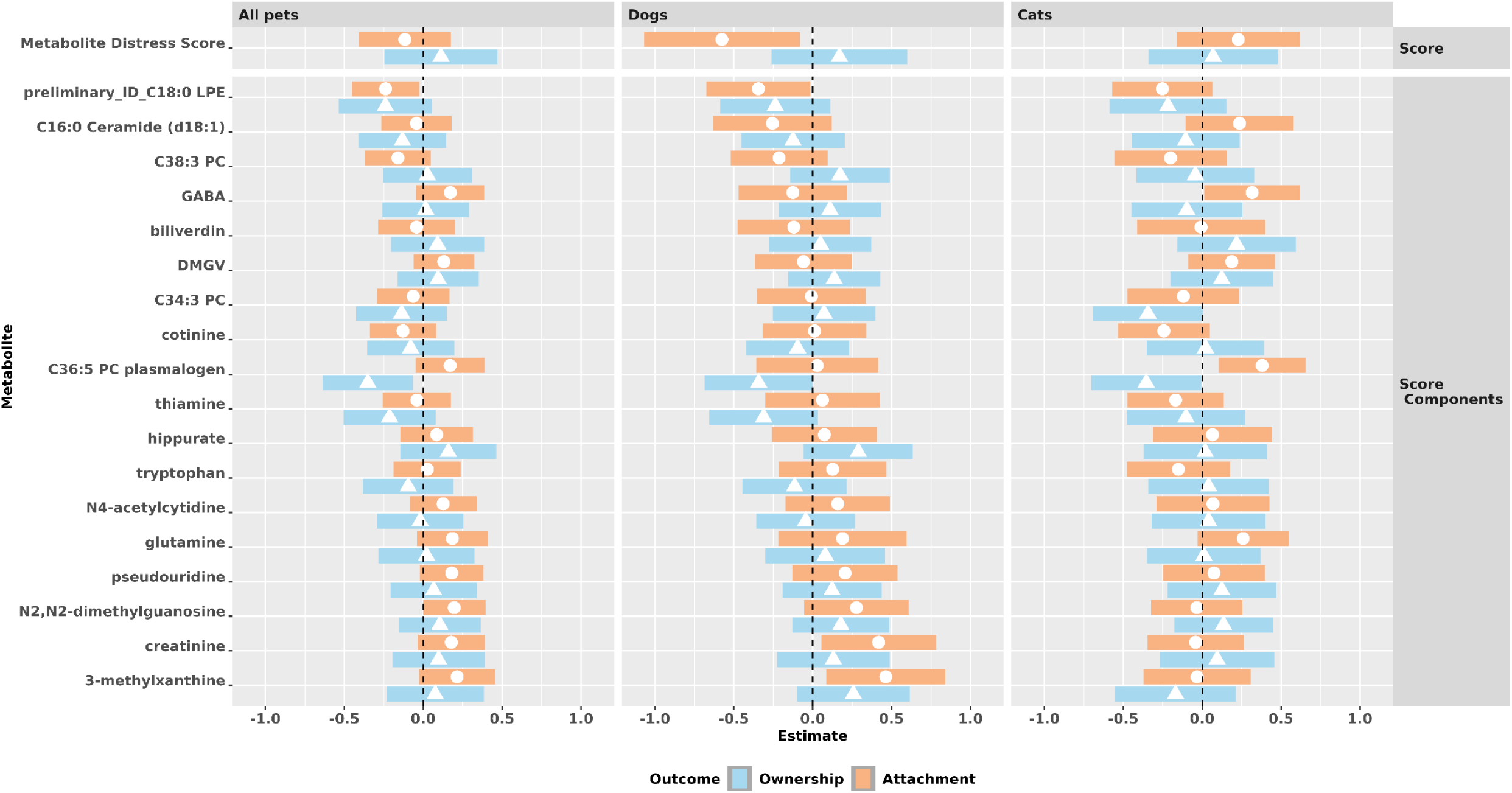
Associations of the metabolite distress score and its constituting individual metabolites with pet ownership and with pet attachment. Results for pet ownership represent mean differences and confidence intervals in transformed metabolite levels comparing pet owners, dog owners or cat owners to those who do not own pets. Results for pet attachment represent mean increases and confidence intervals in transformed metabolite levels for one standard deviation increase in the transformed Lexington Attachment to Pets Scale. Statistical model adjusts of age, date of, fasting status at, and menopausal status at the time of blood collection, race, body mass index, physical activity, Alternative Healthy Eating Index without alcohol, alcohol consumption, subjective sleep quality, and socio-economic status.

**Figure 2.**
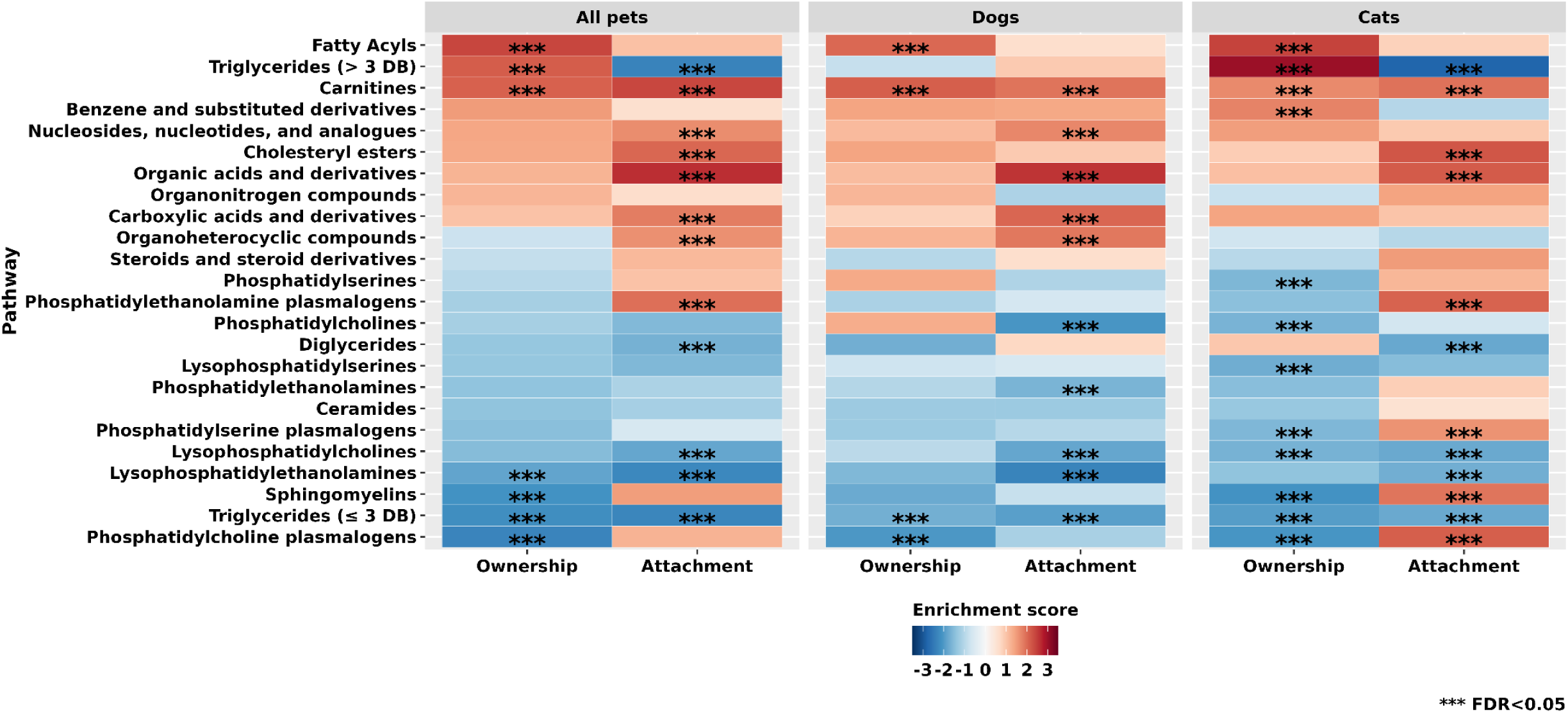
Metabolite classes associated with pet ownership and pet attachment based on Model 3, by pet type. Results represent normalized enrichment scores estimated with Metabolite Set Enrichment Analysis. Positive associations are shown in shades of red while inverse associations are shown in shades of blue. Metabolite classes with FDR<0.05 are marked with ***. Statistical model adjusts of age, date of, fasting status at, and menopausal status at the time of blood collection, race, body mass index, physical activity, Alternative Healthy Eating Index without alcohol, alcohol consumption, subjective sleep quality, and socio-economic status.

Pet attachment among all pet owners (mean increase in MDS levels for 1 standard deviation (SD) increase in pet attachment based on model 3 β (95% CI) = −0.12 (−0.41; 0.17), p = 0.4) and among cat owners (model 3 β (95% CI) = 0.23 (−0.16; 0.62), p = 0.24) were not associated with MDS in either model (Figure 1, Supplementary Figure 2, and Supplementary Table 2). However, pet attachment among dog owners (Figure 1, Supplementary Figure 2, and Supplementary Table 2) was inversely associated with MDS, indicating that stronger attachment to dogs was associated with lower levels of MDS. Effect estimates became stronger with more detailed adjustment for potential confounders and were statistically significant in model 3 (β (95% CI) = −0.58 (−1.07; −0.08), p = 0.02). This association was driven by three metabolites: 3-methylxanthine (β (95% CI) = 0.46 (0.09; 0.84), p = 0.02), creatinine (β (95% CI) = 0.42 (0.06; 0.78; p = 0.02)), and C18:0 lysophosphatidylethanolamine (β (95% CI) = −0.34 (−0.67; −0.02); p = 0.04).

Based on the mediation analysis among dog owners, 13% of the association between pet attachment and a summary measure of multiple self-reported psychological distress measures is mediated by MDS. This finding was not statistically significant (p>0.05) which may be due to the small sample size of N=76 participants who reported owning a dog.

### 3. Metabolomic profiles associated with pet ownership and attachment

#### 3.1 Individual metabolites associated with pet ownership

No individual metabolites were statistically significantly associated with pet ownership after adjusting for testing multiple correlated hypotheses (NEF-p<0.05, Supplementary Figure 3, and Supplementary Table 3). We observed minimal confounding across the three statistical models and will focus on the fully adjusted model 3. Although associations with pet ownership were not statistically significant (p>0.05), triglycerides including mostly saturated and monounsaturated fatty acids showed a tentative positive association with pet ownership in model 2 but an inverse association in model 3 (Supplementary Figure 3 and Supplementary Table 3) which was mostly due to the additional adjustment for physical activity and AHEI. 11 metabolites (mostly plasmalogens and sphingomyelins (SM)) showed lower mean levels among pet owners compared to those not owning pets at a nominal significance level (p<0.05), with a mean difference between the two groups ranging from −0.36 (95% CI = −0.64;-0.08) for C36:5 phosphatidylcholine (PC) plasmalogen to −0.29 (−0.56;-0.02) for beta-alanine. 14 metabolites (mostly fatty acyls, carnitines, and steroids) showed higher mean levels among pet owners compared to those not owning pets at a nominal significance level (p<0.05), with a mean difference between the two groups ranging from 0.30 (95% CI = 0.004;0.59) for 2-aminooctanoic acid to 0.44 (0.14;0.74) for laserpitin. Directions of association for cat and dog ownership were similar to each other and to pet ownership (Supplementary Figure 1 and Supplementary Table 3).

#### 3.2 Metabolite classes associated with pet ownership

Seven metabolite classes were enriched among metabolites associated with pet ownership after accounting for testing multiple correlated hypotheses (FDR<0.05, Supplementary Figure 4, and Supplementary Table 4). Four metabolite classes were enriched among metabolites inversely associated with pet ownership: PC plasmalogens (NES = −2.33, FDR<0.01), TAG with ≤3 double bonds (DB) (NES = −2.18, FDR<0.01), SM (NES = −2.11, FDR<0.01), and lysophosphatidylethanolamines (LPE, NES = - 1.85, FDR<0.01). Three metabolite classes were enriched among metabolites positively associated with pet ownership: fatty acyls (NES = 2.40, FDR<0.01), TAG with >3 DB (NES = 2.13, FDR<0.01), and carnitines (NES = 2.08, FDR<0.01). We observed minimal confounding across the three statistical models except for TAGs with ≤3 DB which were enriched among positively associated metabolites in Model 2 (NES = 2.24, FDR<0.01) and among inversely associated metabolites in Model 3 (NES = −2.18, FDR<0.01), mostly due to the additional adjustment for physical activity and AHEI in Model 3 compared to Model 2. Directions of association for cat and dog ownership were similar to each other and to pet ownership (Figure 1) for most metabolite classes. However, only four metabolite classes were associated with dog ownership at FDR<0.05 (positive associations: carnitines and fatty acyls; inverse associations: TAG with ≤3 DB and PC plasmalogens) while a total of 12 metabolite classes were associated with cat ownership at FDR<0.05 (positive associations: TAG with >3 DB, carnitines, fatty acyls, benzene and substituted derivatives; inverse associations: TAG with ≤3 DB, phosphatidylserines (PS), PC, Lysophosphatidylserines (LPS), PC plasmalogens, LPC, SM, PS plasmalogens).

#### 3.3 Individual metabolites associated with pet attachment

One metabolite, N6,N6-dimethyllysine, was statistically significantly associated with pet attachment (mean increase in metabolite levels for 1 standard deviation (SD) increase in pet attachment in model 3 = 0.39 (95%CI = 0.19;0.59), NEF-p=0.03) after adjusting for testing multiple correlated hypotheses (Supplementary Figure 5 and Supplementary Table 5). We observed minimal confounding across the three statistical models and will focus on the fully adjusted model 3. Twenty-three metabolites (mostly triglycerides (TAG), lysophosphatidylcholines (LPC), and lysophosphatidylethanolamines (LPE)) were inversely associated with pet attachment at a nominal significance level (p<0.05), with a mean increase in metabolite levels per 1 SD increase in pet attachment ranging from −0.29 (95% CI = −0.47;-0.10) for C16:0 LPC to −0.20 (−0.40;-0.004) for C50:5 TAG. Twelve metabolites (mostly organic acids and derivatives) were positively associated with pet attachment at a nominal significance level (p<0.05), with a mean increase in metabolite levels per 1 SD increase in pet attachment from 0.21 (95% CI = 0.01;0.41) for phenylalanine to 0.33 (0.11;0.55) for symmetric dimethylarginine (SDMA). Directions of association for pet attachment among cat and dog owners were similar to those among pet owners (Supplementary Figure 5 and Supplementary Table 5).

#### 3.4 Metabolite classes associated with pet attachment

Twelve metabolite classes were enriched among metabolites associated with pet attachment after accounting for testing multiple correlated hypotheses (FDR<0.05, Supplementary Figure 6 and and Supplementary Table 6). Five metabolite classes were enriched among metabolites inversely associated with pet ownership: TAG with >3DB (NES = −2.35, FDR<0.01), TAG with ≤3 DB (NES = −2.30, FDR<0.01), LPE (NES = −2.26, FDR<0.01), LPC (NES = −1.82, FDR<0.01), and diglycerides (NES = −1.67, FDR=0.01). Seven metabolite classes were enriched among metabolites positively associated with pet ownership: organic acids and derivatives (NES = 2.63, FDR<0.01), carnitines (NES = 2.38, FDR<0.01), cholesteryl esters (CE, NES = 2.02, FDR<0.01), phosphatidylethanolamines (PE) plasmalogens (NES = 1.95, FDR<0.01), carboxylic acids and derivatives (NES = 1.82, FDR<0.01), nucleosides, nucleotides, and analogues (NES = 1.65, FDR<0.01), and organoheterocyclic compounds (NES = 1.62, FDR = 0.02). Pattern of associations with pet attachment varied between dog and cat owners (Figure 1). CE, PE plasmalogens, PC plasmalogens, SM, PS plasmalogens, DAG, and TAG with >3 DB were associated with pet attachment among cat owners but not among dog owners while organoheterocyclic compounds, nucleosides, nucleotides, and analogues, carboxylic acids and derivatives, PC, and PE were associated with pet attachment among dog owners but not among cat owners.

#### 3.5 Comparison of pet ownership with pet attachment

Most metabolite classes showed similar associations with pet ownership and pet attachment among all pet owners and among dog owners (Figure 3, Supplementary Figures 5 and 6). However, TAG with >3DB were positively associated with pet ownership (NES=2.13, FDR<0.05) but inversely associated with pet attachment (NES= −2.35, FDR<0.05) among all pet owners.

Multiple metabolite classes showed different directions of association with cat ownership compared to cat attachment (Figure 3, Supplementary Figures 5 and 6). TAG with >3DB were positively associated with pet ownership (NES=3.01, FDR<0.05) but inversely associated with pet attachment (NES=-2.08, FDR<0.05) among cat owners. Phosphatidylserine (PS) plasmalogens, sphingomyelins, and PC plasmalogens were inversely associated with pet ownership (NES=-1.59; NES =-2.11; NES=-2.07; all FDR<0.05) but positively associated with pet attachment (NES=1.59; NES=1.90; NES=2.11; all FDR<0.05) among cat owners.

## Discussion

We conducted the first study to assess the association of pet ownership and attachment with psychological distress-related metabolic dysregulation. We estimated psychological distress-related metabolic dysregulation with a metabolite-based distress score (MDS) previously developed in this cohort and found no statistically significant associations between pet, dog or cat ownership and MDS. Similarly, we found no statistically significant association between pet attachment among all pet owners or among cat owners and MDS. However, we found a statistically significant inverse association between pet attachment among dog owners and MDS. We also identified several metabolite classes that are statistically significantly associated with pet ownership and pet attachment.

### MDS and pet ownership and attachment

In our study, we observed an inverse association between pet attachment among dog owners and MDS suggesting that higher attachment to one’s companion dog is associated with metabolomic profiles reflecting lower psychological distress. Psychological distress, including depression and anxiety, represents a public health burden and is more common among women. Psychological distress has been associated with multiple common chronic diseases, including diabetes, cardiovascular disease and cancer [23–25], and proposed mechanisms include increased inflammation and dysregulation of the endocrine system, the hypothalamic–pituitary–adrenal axis and the immune system [26]. Findings from our study suggest that metabolic dysregulation represents a potential novel biological mechanism contributing to this association. Although statistically not significant, results from our mediation analysis suggest that 13% of the proportion of the association between a summary measure of multiple self-reported psychological distress measures and pet attachment is mediated by psychological distress-related metabolic-dysregulation estimated by MDS. MDS has been derived by our group in the Nurses’ Health Study and we have showed that MDS is positively associated with cardiovascular disease risk in two independent cohort studies, the Women’s Health Initiative Observational Study (WHI-OS) and the Spanish Prevencion con Dieta Mediterranea (PREDIMED) trial [11], which includes both men and women. If our results are replicated in other cohorts and prospective studies, the addition of a dog to a person’s life, if that person is likely to form a high attachment to their dog, may provide a novel intervention strategy to mitigate the negative metabolic effects of psychological distress and potentially offer protection from devolving chronic illnesses such as cardiovascular disease. Additional studies are necessary to assess the association of MDS with other chronic illnesses.

The association between dog attachment and MDS was driven by three individual metabolites: creatinine and 3-methylxanthine were positively associated with pet attachment while and C18:0 lysophosphatidylethanolamine (LPE) was inversely associated with pet attachment. Creatinine is a nitrogenous organic acid and the end product of creatine and creatine phosphate metabolism [27]. Serum creatinine is produced in muscle cells and represents the most widely used biomarker of kidney function [28]. Because of its production in muscle cells, serum creatinine has also been proposed as an estimate of muscle mass [29, 30]. In line with several previous studies indicating dog ownership and attachment is associated with high levels of physical activity [31–34], higher levels of creatinine were associated with high levels of pet attachment in our study. While our analysis included adjustment for physical activity, BMI and other lifestyle factors, it is possible that this association is due to residual confounding by these factors as they are all associated with muscle mass. However, animal model studies suggest that plasma creatinine levels may reflect creatine levels in the brain [35] and that alterations in the creatine system may play a role in the development of depressive symptoms [36, 37]. Furthermore, creatine supplementation has been suggested to have antidepressants effects [35]. 3-methylxanthine was positively associated with pet attachment in our study. 3-methylxanthine is a methylxanthine, a group of caffeine-related metabolites with anti-inflammatory properties [38] and the ability to release catecholamines, including dopamine and norepinephrine, from the adrenal gland [39, 40]. The release of catecholamines promotes stress-induced adaptation and thus, may have antidepressant effects. LPEs are the second-most abundant lysoglycerophospholipids with high concentrations in interstitial fluids and plasma. LPEs are generated by PLA2 from phosphatidylethanolamines (PE) in cell membranes. The blood-brain barrier transporter Mfsd2a can bind to LPE, suggesting that LPEs can cross the blood-brain-barrier from plasma [41]. Pet attachment was inversely associated with C18:0 LPE in our study. Importantly, human studies consistently observed increased LPE levels among people with depression [42, 43] and anxiety (LPE C18:2/0:0) [44]. If our findings are replicated in other independent prospective cohort studies, our study suggests that higher attachment to dogs but not cats is associated with higher creatine and 3-methylxanthine levels and with lower LPE levels which all may have antidepressant effects.

### Metabolite classes and pet ownership and attachment

Several more metabolite classes were associated with pet ownership and pet attachment. Most directions of associations were similar for pet ownership and pet attachment but we observed multiple different directions of associations when comparing cat ownership with cat attachment.

### Triglycerides

Triglycerides (TAG) represent a key component of fatty acid metabolism, which was shown to be altered in depression. A recent meta-analysis of nine Dutch cohorts, including 10,145 control subjects and 5,283 persons with depression established with diagnostic interviews or questionnaires, found that higher levels of TAGs were associated with higher odds of depression [45] while another small human study of major depressive disorder found that only specific TAGs, TAGs with >4 DBs, were positively associated with depression [46]. In our study, TAGs with ≤3 DB were inversely associated with both pet ownership and pet attachment, among dogs and cat owners. Interestingly TAGs with >3 DB were positively associated with pet ownership and inversely associated with pet attachment, which was driven by differences among cat owners. Our findings, if replicated, suggest that attachment to cats but not to dogs may lower levels of TAGs with >3 DB and potentially mitigate some of the psychological distress-related lipid dysregulation.

### Sphingomyelins

Sphingomyelins (SMs) are the most abundant class of sphingolipids, essential for cell membrane and cell function. SM are synthesized from ceramides and hydrolyzed back into ceramides, they play an important role in cell signaling and the immune system and have anti-inflammatory properties [47–54]. The research on sphingomyelins (SM) and psychological distress is scarce. A Dutch family-based study found a significant inverse correlation between multiple SM measures (SM 23:1/SM 16:0, SM 23:1/SMs, SMs) and multiple measures of depression and anxiety (major depressive disorder, Hospital Anxiety and Depression Scales HADS-D, HADS-A and the Centre for Epidemiological Studies Depression Scale CES-D) [55, 56]. A study examining the brain lipidome of rats found that many SMs were negatively correlated with stress [57]. In our study, sphingomyelins were inversely associated with pet and cat ownership but positively associated with cat attachment. Additional studies are required to assess the association of SM with psychological distress and any potential role that pet ownership and attachment might play.

### Phosphatidylcholine plasmalogens (PCP)

Plasmalogens are common glycerophospholipids that rigidify membranes and can scavenge free radicals. Phosphatidylcholine plasmalogens (PCP) are formed via headgroup transfer from phosphatidylethanolamine plasmalogens [58]. In our study, PCPs were inversely associated with pet and cat ownership but positively associated with cat attachment. In a previous study, our group has found C36:5 PCP to be inversely associated with chronic distress among middle-age and older women [11] but no other studies assessed the association of PCPs with psychological distress. Plasmalogens increase early in life and then rapidly decrease with increasing age and they may play a role in neurodegenerative disorders [58]. Additional studies are required to assess the association of PCPs with psychological distress and any potential role that pet ownership and attachment might play.

Our study has several strengths and limitations. Although cross-sectional by design, this is the first study to assess the association of psychological distress-related metabolic dysregulation with pet ownership and attachment and to identify metabolic profiles associated with these exposures.

Additional strengths include the availability of a wide range of metabolites, detailed covariate information and careful adjustment for potential confounders. Our cohort consisted of predominantly White registered nurses, a group who is not representative of the general population. While we found several statistically significant associations, our sample size was limited in the analyses stratified by pet type. However, we applied stringed QC criteria to limit the identification of spurious associations.

Another limitation is that we only analyzed blood samples collected at one point in time; however, we previously showed that the majority of the measured metabolites have a high within person stability over 1-2 and over 10 years [19, 20]. Further, we do not have an independent validation dataset. As this type of data becomes more common in human-animal interaction studies, further population studies are needed to validate the results discussed here, while experimental studies are required to understand the biological mechanisms underlying these associations.

In summary, although MDS was not associated with pet ownership or attachment, we observed an inverse association between MDS and pet attachment among dog owners, but not among cat owners. Additionally, several metabolite groups were associated with pet ownership and attachment. Most associations were similar in directions among dog owners while some differences were observed when comparing cat ownership to cat attachment.

### Data availability statement

Due to participant confidentiality and privacy concerns, data cannot be shared publicly and requests to access NHS/NHSII data must be submitted in writing. According to standard controlled access procedures, applications to use NHS/NHSII resources will be reviewed by our External Collaborations Committee to verify that the proposed use maintains the protection of the privacy of participants and the confidentiality of the data. Investigators wishing to use NHS/NHSII data are asked to submit a brief description of the proposed project. Please see https://www.nurseshealthstudy.org/researchers (contact) for details.

## Funding

This study was supported by the Eunice Kennedy Shriver National Institute of Child Health and Human Development (grant No. R01 HD101101; PI: Dr Schernhammer) as well as National Institutes of Health (U01 HL145386, U01 CA176726, U01 CA167552). The content is solely the responsibility of the authors and does not necessarily represent the official views of the National Institutes of Health.

## Author Contributions

ESS conceived the study. OAZ, SS and ESS designed the study. OAZ performed the statistical analyses and drafted the manuscript. All authors interpreted the results, critically revised the manuscript and approved the final version.

## Declaration of interests

The authors declare no competing interests.

## Notes

### Competing Interest Statement

The authors have declared no competing interest.

### Author Declarations

The study protocol was approved by the Institutional Review Board of Brigham and Women's Hospital and the Committee on the Use of Human Subjects in Research of Harvard T.H. Chan School of Public Health (Boston, MA, USA). Voluntary return of questionnaires indicates informed consent. The study was conducted in accordance with all relevant ethical guidelines and regulations, including the Declaration of Helsinki.

