## Supplementary Figures for "Associations of pet ownership and pet attachment with a metabolite-based psychosocial distress score and circulating metabolomic profiles"

**Supplementary Figure 1. Associations between pet ownership and the metabolite distress score, including its constituting individual metabolites.** Results represent mean differences in transformed metabolite levels comparing pet owners, dog owners or cat owners to those who do not own pets. Positive associations are shown in shades of red while inverse associations are shown in shades of blue. Statistical significance is overlaid on the plot: *p<0.05, ** Number of effective tests corrected (NEF) p<0.2, *** NEF-p<0.05. Model 1 adjusts of age. Model 2 additionally adjusts for date of, fasting status at, and menopausal status at the time of blood collection, and race. Model 3 additionally adjusts for body mass index, physical activity, Alternative Healthy Eating Index without alcohol, alcohol consumption, subjective sleep quality, and socio-economic status.

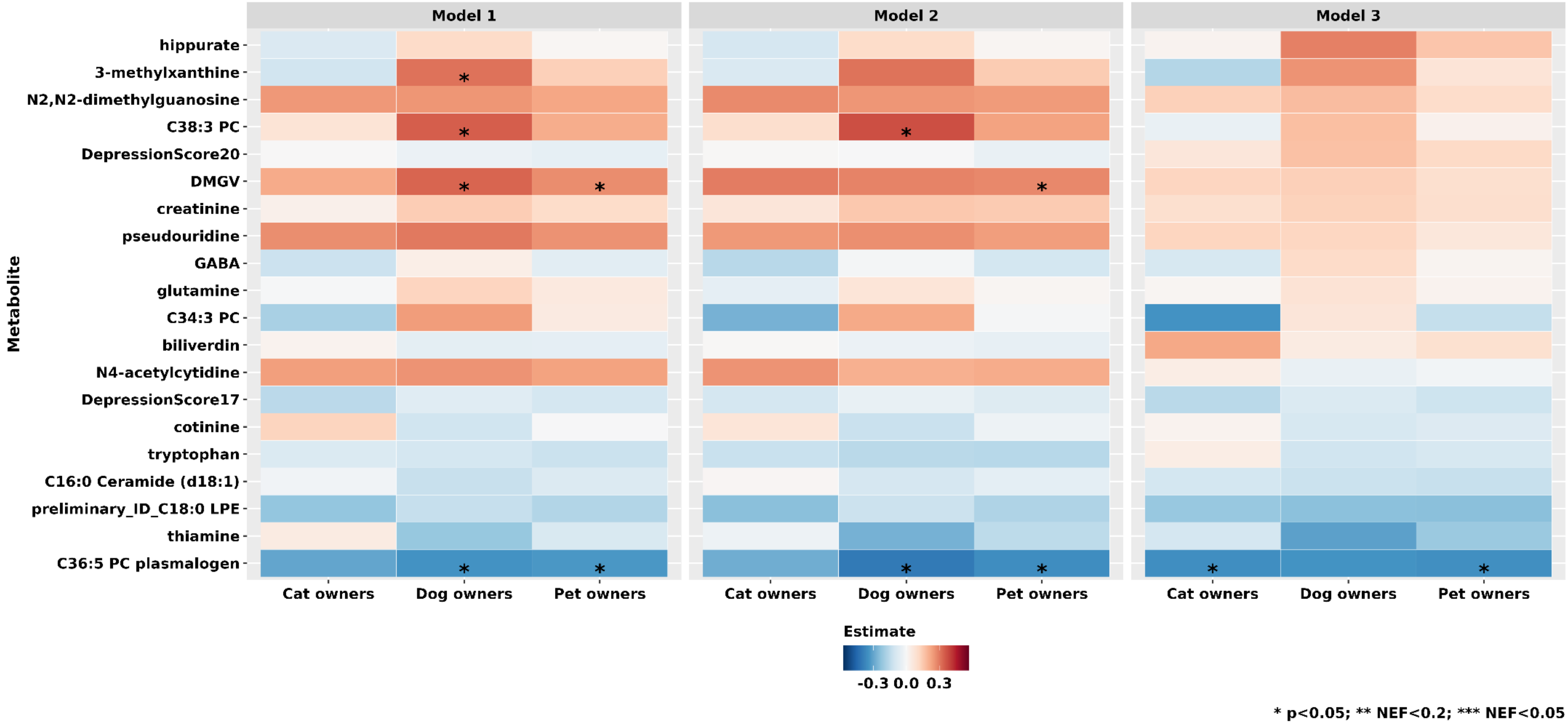

**Supplementary Figure 2. Associations between pet attachment and the metabolite distress score, including its constituting individual metabolites.** Results represent mean increase in transformed metabolite levels for one standard deviation increase in the transformed Lexington Attachment to Pets Scale. Positive associations are shown in shades of red while inverse associations are shown in shades of blue. Statistical significance is overlaid on the plot: *p<0.05, ** Number of effective tests corrected (NEF) p<0.2, *** NEF-p<0.05. Model 1 adjusts of age. Model 2 additionally adjusts for date of, fasting status at, and menopausal status at the time of blood collection, and race. Model 3 additionally adjusts for body mass index, physical activity, Alternative Healthy Eating Index without alcohol, alcohol consumption, subjective sleep quality, and socio-economic status.

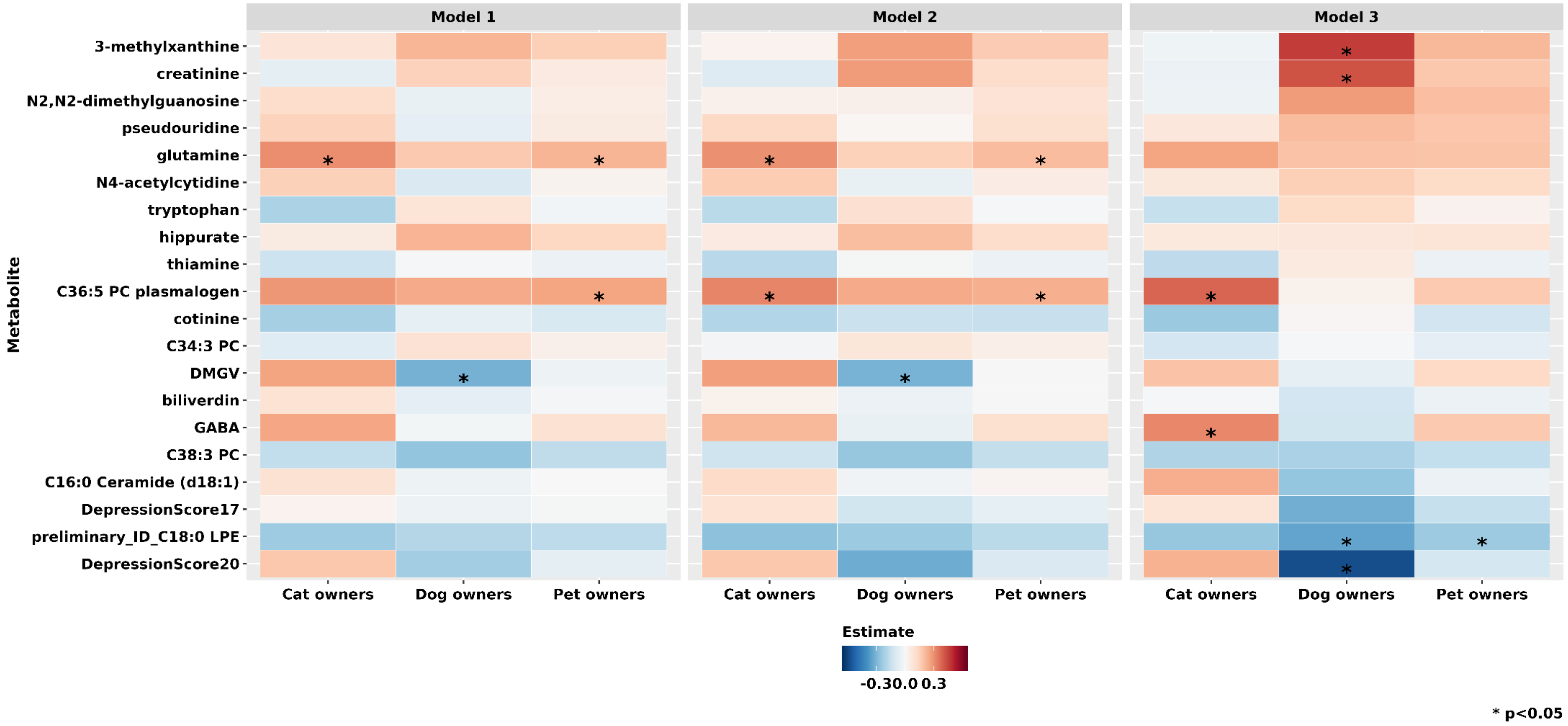

**Supplementary Figure 3. Metabolites associated with pet ownership, cat ownership and dog ownership.** Results represent mean differences in transformed metabolite levels comparing pet owners or dog owners or cat owners to those who do not own pets. Positive associations are shown in shades of red while inverse associations are shown in shades of blue. Statistical significance is overlaid on the plot: *p<0.05, ** Number of effective tests corrected (NEF) p<0.2, *** NEF-p<0.05. Model 1 adjusts of age. Model 2 additionally adjusts for date of, fasting status at, and menopausal status at the time of blood collection, and race. Model 3 additionally adjusts for body mass index, physical activity, Alternative Healthy Eating Index without alcohol, alcohol consumption, subjective sleep quality, and socio-economic status.

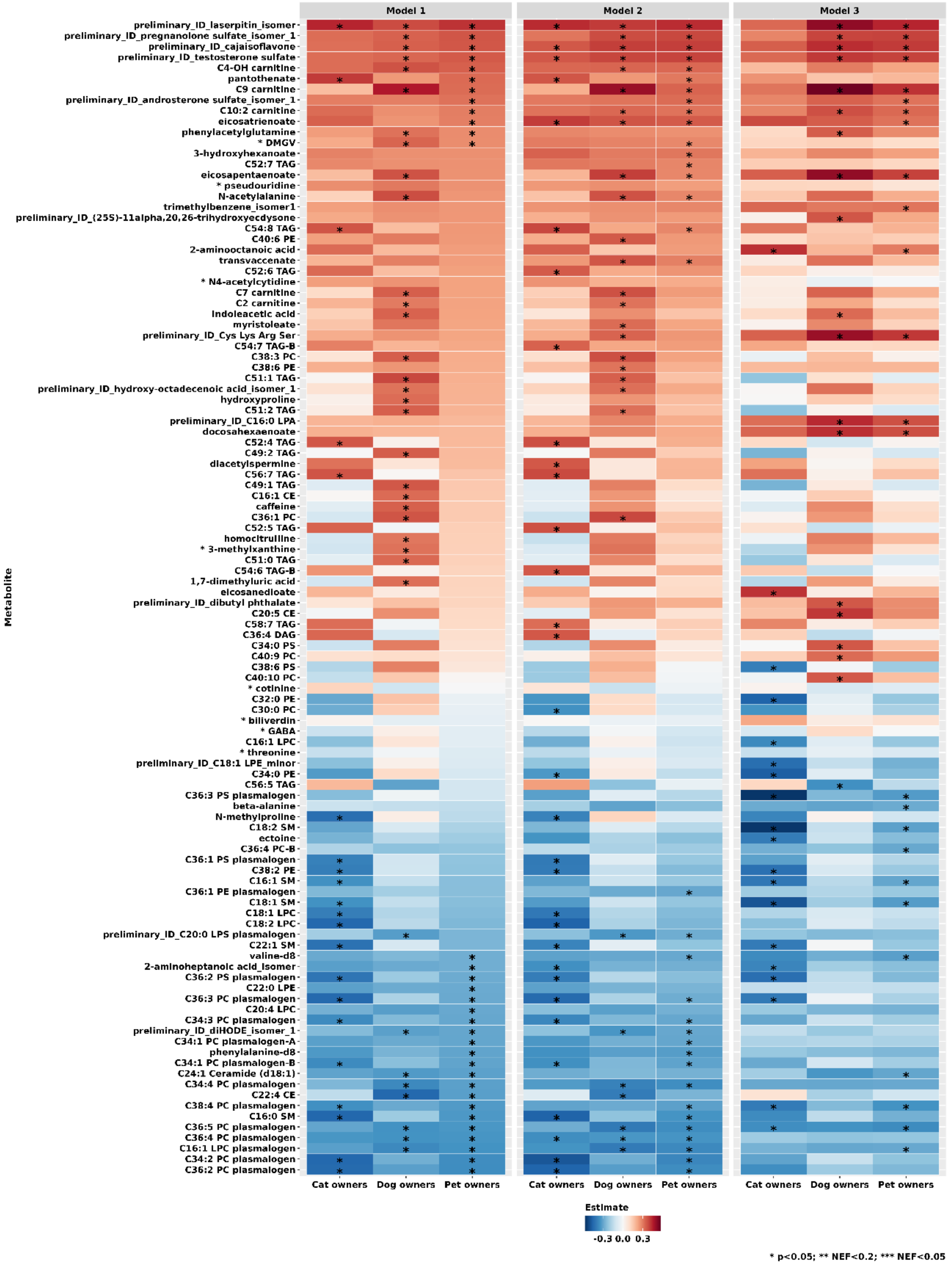

**Supplementary Figure 5. Metabolites associated with pet attachment among all pet owners, among cat owners, and among dog owners.** Results represent mean increase in transformed metabolite levels for one standard deviation increase in the transformed Lexington Attachment to Pets Scale. Positive associations are shown in shades of red while inverse associations are shown in shades of blue. Statistical significance is overlaid on the plot: *p<0.05, ** Number of effective tests corrected (NEF) p<0.2, *** NEF-p<0.05. Model 1 adjusts of age. Model 2 additionally adjusts for date of, fasting status at, and menopausal status at the time of blood collection, and race. Model 3 additionally adjusts for body mass index, physical activity, Alternative Healthy Eating Index without alcohol, alcohol consumption, subjective sleep quality, and socio-economic status.

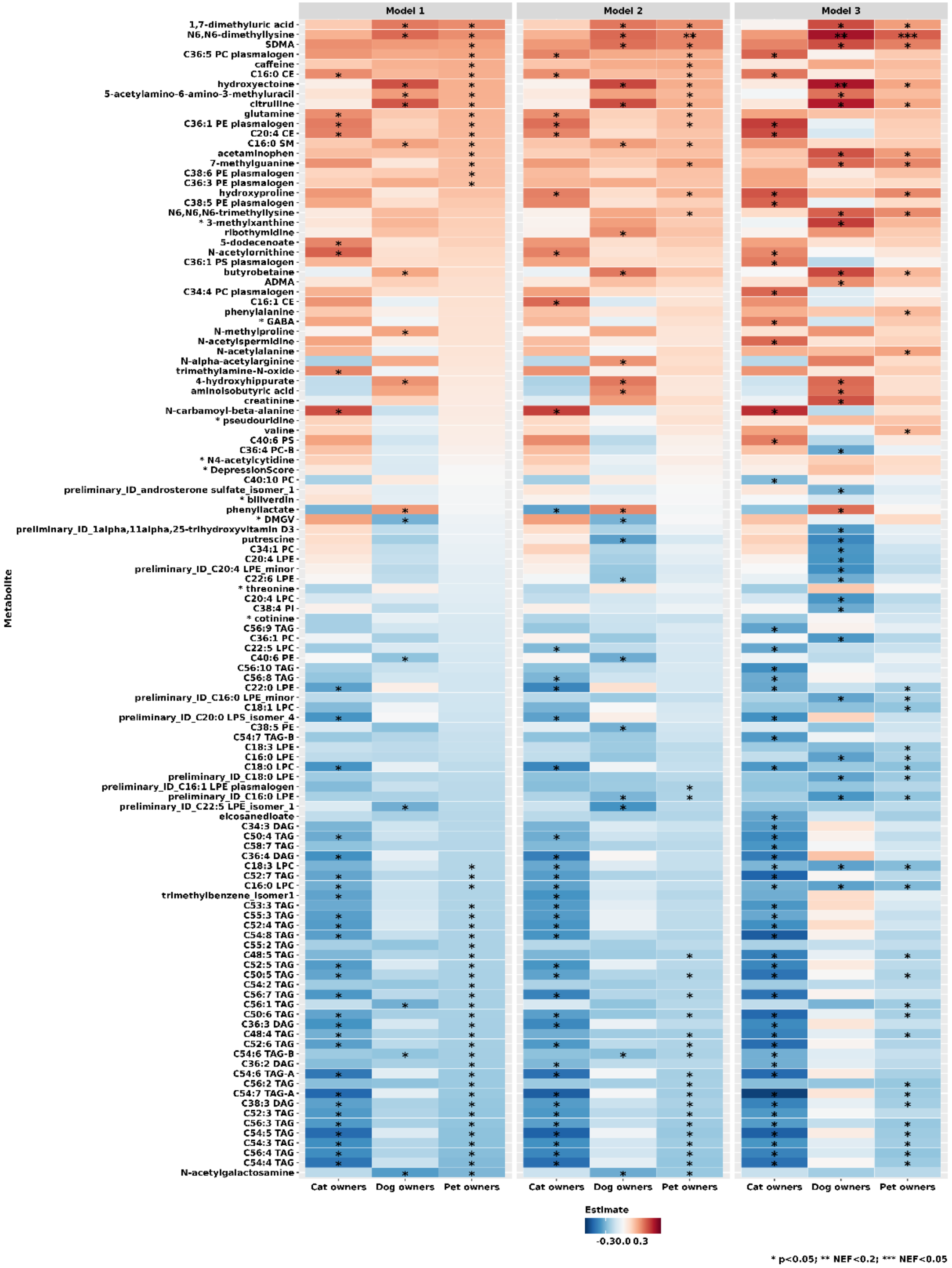

**Supplementary Figure 4. Metabolite classes associated with pet ownership, cat ownership and dog ownership.** Results represent normalized enrichment scores estimated with Metabolite Set Enrichment Analysis. Positive associations are shown in shades of red while inverse associations are shown in shades of blue. Metabolite classes with FDR<0.005 are marked with ***. Model 1 adjusts of age. Model 2 additionally adjusts for date of, fasting status at, and menopausal status at the time of blood collection, and race. Model 3 additionally adjusts for body mass index, physical activity, Alternative Healthy Eating Index without alcohol, alcohol consumption, subjective sleep quality, and socio-economic status.

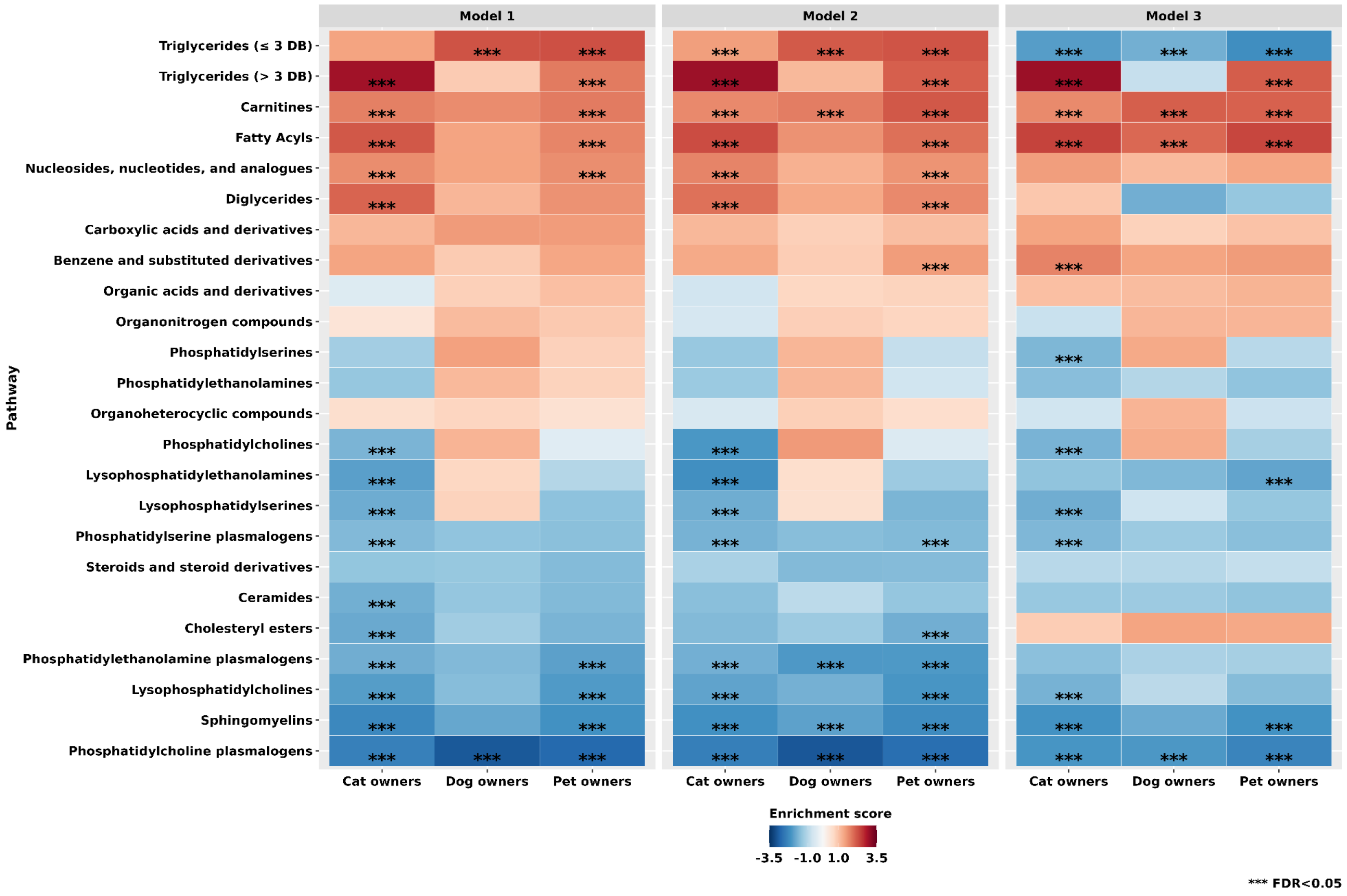

**Supplementary Figure 6. Metabolite classes associated with pet attachment among all pet owners, among cat owners, and among dog owners.** Results represent normalized enrichment scores estimated with Metabolite Set Enrichment Analysis. Positive associations are shown in shades of red while inverse associations are shown in shades of blue. Metabolite classes with FDR<0.005 are marked with ***. Model 1 adjusts of age. Model 2 additionally adjusts for date of, fasting status at, and menopausal status at the time of blood collection, and race. Model 3 additionally adjusts for body mass index, physical activity, Alternative Healthy Eating Index without alcohol, alcohol consumption, subjective sleep quality, and socio-economic status.

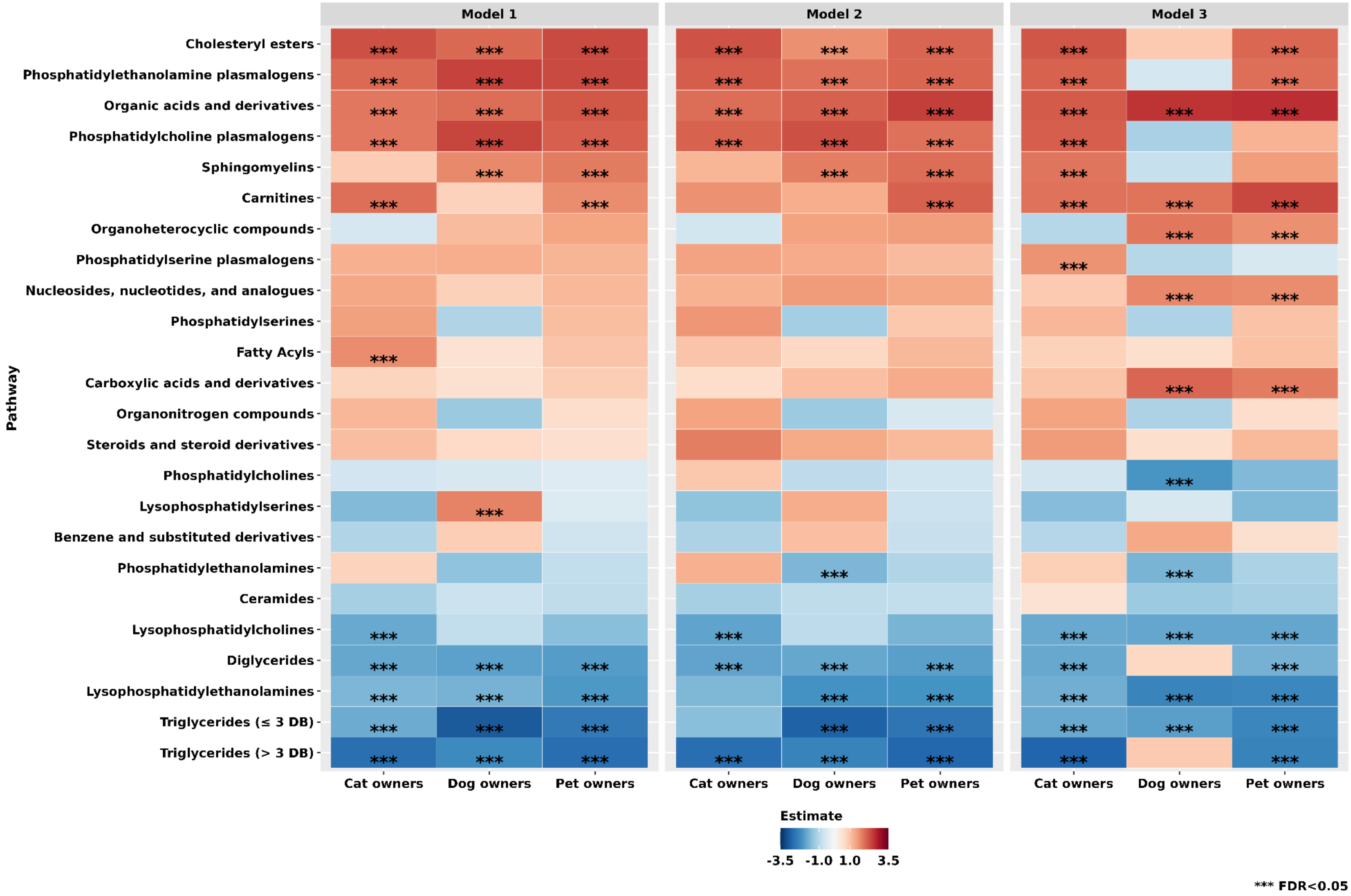
