## Supplementary Methods for "Associations of pet ownership and pet attachment with a metabolite-based psychosocial distress score and circulating metabolomic profiles"

**Supplementary materials**

**Metabolite profiling**

Plasma metabolites were profiled at the Broad Institute of MIT and Harvard (Cambridge, MA) using three complementary liquid chromatography tandem mass spectrometry (LC-MS/MS) methods designed to measure polar metabolites and lipids as well as free fatty acids as described previously [16-19]. Additional details are presented in the supplementary materials. For each method, pooled plasma reference samples were included every 20 samples and results were standardized using the ratio of the value of the sample to the value of the nearest pooled reference multiplied by the median of all reference values for the metabolite. In addition, 64 quality control (QC) samples, to which the laboratory was blinded, were also profiled. These were randomly distributed among the participants' samples.

Hydrophilic interaction liquid chromatography (HILIC) analyses of water-soluble metabolites in the positive ionization mode were conducted using an LC-MS system comprised of a Shimadzu Nexera X2 U-HPLC (Shimadzu Corp.; Marlborough, MA) coupled to a Q Exactive mass spectrometer (Thermo Fisher Scientific; Waltham, MA). Metabolites were extracted from plasma (10 µL) using 90 µL of acetonitrile/methanol/formic acid (74.9:24.9:0.2 v/v/v) containing stable isotope-labeled internal standards (valine-d8, Sigma-Aldrich; St. Louis, MO; and phenylalanine-d8, Cambridge Isotope Laboratories; Andover, MA). The samples were centrifuged (10 min, 9,000 x g, 4°C), and the supernatants were injected directly onto a 150 x 2 mm, 3 µm Atlantis HILIC column (Waters; Milford, MA). The column was eluted isocratically at a flow rate of 250 µL/min with 5% mobile phase A (10 mM ammonium formate and 0.1% formic acid in water) for 0.5 minute followed by a linear gradient to 40% mobile phase B (acetonitrile with 0.1% formic acid) over 10 minutes. MS analyses were carried out using electrospray ionization in the positive ion mode using full scan analysis over 70-800 m/z at 70,000 resolution and 3 Hz data acquisition rate. Other MS settings were: sheath gas 40, sweep gas 2, spray voltage 3.5 kV, capillary temperature 350°C, S-lens RF 40, heater temperature 300°C, microscans 1, automatic gain control target 1e6, and maximum ion time 250 ms.

Plasma lipids were profiled using a Shimadzu Nexera X2 U-HPLC (Shimadzu Corp.; Marlborough, MA). Lipids were extracted from plasma (10 µL) using 190 µL of isopropanol containing 1,2-didodecanoyl-sn-glycero-3-phosphocholine (Avanti Polar Lipids; Alabaster, AL). After centrifugation, supernatants were injected directly onto a 100 x 2.1 mm, 1.7 µm ACQUITY BEH C8 column (Waters; Milford, MA). The column was eluted isocratically with 80% mobile phase A (95:5:0.1 vol/vol/vol 10mM ammonium acetate/methanol/formic acid) for 1 minute followed by a linear gradient to 80% mobile-phase B (99.9:0.1 vol/vol methanol/formic acid) over 2 minutes, a linear gradient to 100% mobile phase B over 7 minutes, then 3 minutes at 100% mobile-phase B. MS analyses were carried out using electrospray ionization in the positive ion mode using full scan analysis over 200–1100 m/z at 70,000 resolution and 3 Hz data acquisition rate. Other MS settings were: sheath gas 50, in source CID 5 eV, sweep gas 5, spray voltage 3 kV, capillary temperature 300°C, S-lens RF 60, heater temperature 300°C, microscans 1, automatic gain control target 1e6, and maximum ion time 100 ms. Lipid identities were denoted by total acyl carbon number and total double bond number.

Metabolites of intermediate polarity, including free fatty acids and bile acids, were profiled using a Nexera X2 U-HPLC (Shimadzu Corp.; Marlborough, MA) coupled to a Q Exactive (Thermo Fisher Scientific; Waltham, MA). Plasma samples (30 µL) were extracted using 90 µL of methanol containing PGE2-d4 as an internal standard (Cayman Chemical Co.; Ann Arbor, MI) and centrifuged (10 min, 9,000 x g, 4°C). The supernatants (10 µL) were injected onto a 150 x 2.1 mm ACQUITY BEH C18 column (Waters; Milford, MA). The column was eluted isocratically at a flow rate of 450 µL/min with 20% mobile phase A (0.01% formic acid in water) for 3 minutes followed by a linear gradient to 100% mobile phase B (0.01% acetic acid in acetonitril) over 12 minutes. MS analyses were carried out using electrospray ionization in the negative ion mode using full scan analysis over m/z 70-850. Additional MS settings are: ion spray voltage, -3.5 kV; capillary temperature, 320°C; probe heater temperature, 300 °C; sheath gas, 45; auxiliary gas, 10; and S-lens RF level 60.

Raw data from orbitrap mass spectrometers were processed using TraceFinder 3.3 software (Thermo Fisher Scientific; Waltham, MA) and Progenesis QI (Nonlinear Dynamics; Newcastle upon Tyne, UK) and targeted data from the QTRAP 5500 system were processed using MultiQuant (version 2.1, SCIEX; Framingham, MA). For each method, metabolite identities were confirmed using authentic reference standards or reference samples.
